# Physician Engagement in Quality Improvement: A Cross-Sectional Pilot Study

**DOI:** 10.1101/2023.04.27.23289231

**Authors:** Christine Shea, Laure Perrier, Melissa Prokopy, Monique Herbert, Sundeep Sodhi, Alia Karsan, Julie Simard, Tyrone A. Perreira

**Author notes:** Correspondence: Christine Shea. Dalla Lana School of Public Health, Institute of Health Policy, Management and Evaluation, University of Toronto, 155 College Street, Suite 425, Toronto M5T 3M6, Ontario, Canada. **Authors’ contributions** TP, CS, MP, MH, JS conceived of the idea. TP, CS, MP conducted the pilot study. MH and TP were responsible for data analysis. CS wrote the manuscript and all authors provided editorial advice. All authors read and approved the final manuscript.

## Abstract

**Background:** To confirm the reliability of a survey investigating physician engagement in quality improvement (QI) among Ontario physicians. We conducted a pilot study to test the survey on physicians, evaluate a recruitment strategy, and assess preliminary data.

**Methods:** All Ontario physicians were invited to participate in the survey through province-wide online physician and hospital organization newsletters.

**Results:** Results indicate a need for solutions and standards for training physicians interested in participating in QI initiatives. Study objectives were reached, but recruitment remains challenging.

**Conclusion:** This pilot study supports conducting a full-scale survey that would result in more robust results.

## INTRODUCTION

Consistent delivery of high-quality care remains a never-ending challenge in the face of continuous technical and clinical innovations, rising costs, and an ever-changing system [1]. The quality of care delivered by the health care system rests on the smooth running of a complex network of processes and pathways that must be delivered by people working together harmoniously [2]. When health care processes and pathways do not function optimally, quality improvement (QI) methods and tools used systematically results in tangible, measurable improvements [2]. Physicians understand the interrelated demands of the healthcare system and patient needs, making them well positioned to participate in QI initiatives and address challenges to providing optimal care [3].

Physician involvement in QI has been linked to the success and sustainability of improvement initiatives [4-6] despite a variety of challenges to physician participation in QI work. Challenges include countering traditional expectations regarding physician roles that emphasize a focus on care at the patient-level rather than the system-level [7], a longstanding culture of autonomy [8] and a lack of physician inclusion in the development of organisational policies, processes and systems despite physicians feeling that their inclusion would improve the quality of patient care and their own professional fulfillment [9]. This research indicates that it is necessary to engage with physicians at all points in the system to engage them in health care system improvement.

The potential for meaningful and sustainable improvements to the quality of care is contingent on the ability of those working in this system to understand, accurately evaluate, and intervene appropriately [10]. Physicians at the point of care are optimally positioned for this work. An informed overview of physicians’ interests in QI, opportunities to be involved in QI efforts, and insights into physicians’ experiences of participation, both in hospital and general practice is critical to understand the challenges and opportunities for physician engagement in QI. We developed a survey to support the accurate evaluation of physician engagement in QI. We confirmed the reliability of the questions with a group of hospital physicians. The purpose of the current pilot study is to test the feasibility of the survey and provide preliminary data to refine the processes for conducting a future large-scale survey. Our objectives were to: 1) determine if the survey instrument can be completed by physicians practicing outside of hospital settings; 2) assess a recruitment strategy; and 3) assess preliminary data.

## METHODS

### Study design, sample, and survey administration

A cross-sectional survey was designed to evaluate physician engagement in QI and survey development has been described in a previous publication (Perreira 2020). In brief, we developed the survey by first conducting a series of focused literature searches. We then assembled a group of QI experts to participate in a modified Delphi panel using a convenience sample of physicians. Cognitive debriefing was conducted, and we confirmed the reliability of the questions [11]. For this study, we define QI as, “the combined and unceasing efforts of everyone—healthcare professionals, patients and their families, researchers, payers, planners and educators—to make the changes that will lead to better patient outcomes (health), better system performance (care) and better professional development” [3].

All Ontario physicians were eligible to participate in the survey. Physicians were recruited into the study through two newsletters. The OMA (Ontario Medical Association) and OHA (Ontario Hospital Association) newsletters listed an invitation to participate and a link to the online survey. The OMA newsletter was sent directly to physician members. The OHA newsletter was disseminated using existing distribution lists of senior hospital employees who were then asked to distribute directly to their physicians. A reminder was provided two weeks later.

### Data collection

Participants accessed the online survey with a link. The survey was administered through Checkbox (Checkbox Survey Solutions Inc, USA) in November 2021. The study was approved by the University of Toronto Research Ethics Board (RIS Human Protocol Number 40771), and consent was obtained from each participant. Data was imported into Excel (Microsoft, USA) from the online survey tool.

### Statistical analyses

Statistical analysis was completed with R (R Core Team, Austria) and descriptive analyses were conducted. Frequency distributions were generated for each variable. The relationship between years of practice and receipt of QI training and the relationship between physician gender, receipt of QI training, and interest in receiving QI training were analyzed.

## RESULTS

A total of 231 Ontario licensed physicians completed the study (Table 1). Fifty-three percent of participants (121 out of 231) practiced outside of hospitals. More than half of the group (52%) averaged 22 clinical days or more per month, and 43% had been practicing for 20 years or more. Just over half the group was male (52%). Physician specialty is reported in S1 Appendix.

**Table 1.**
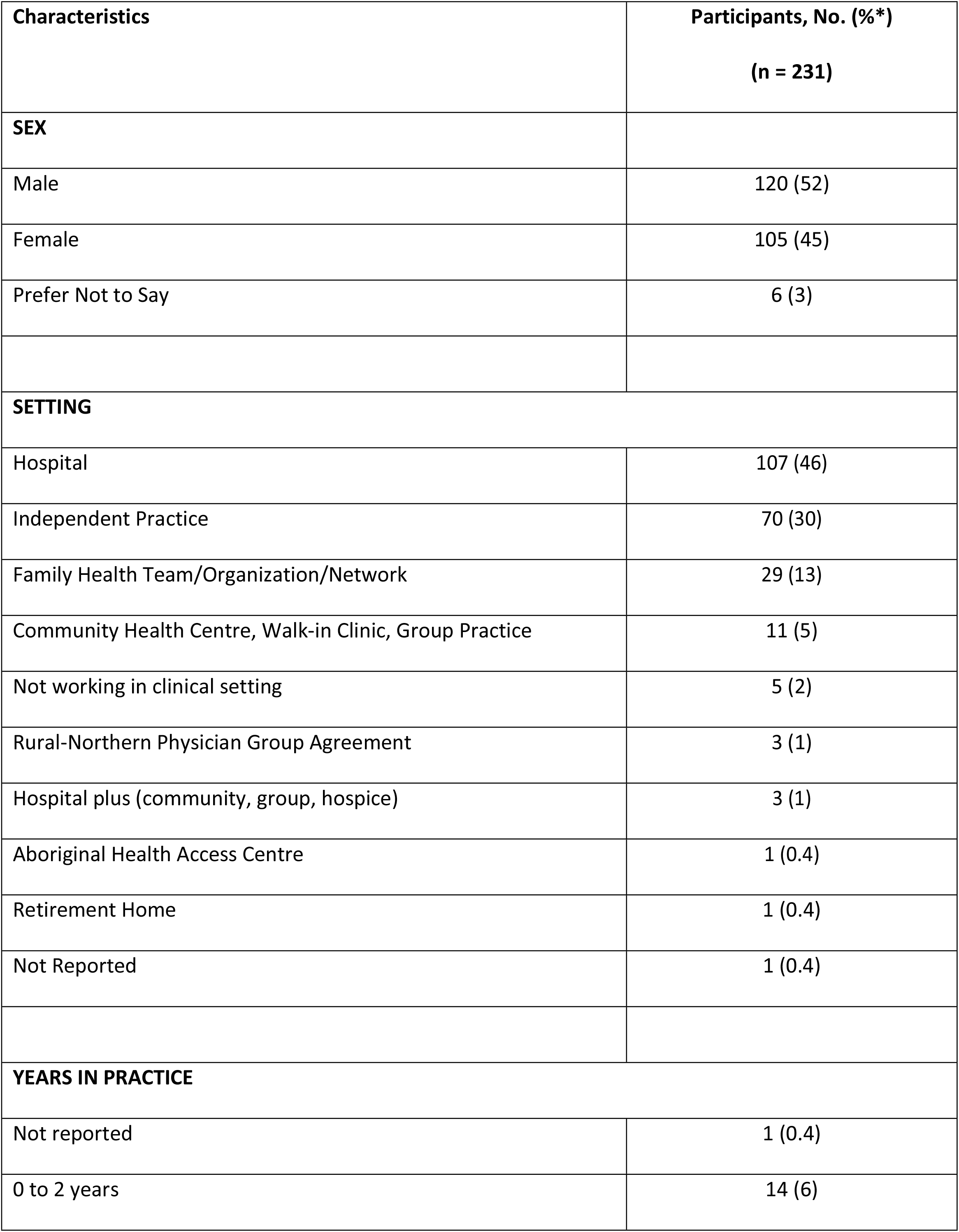
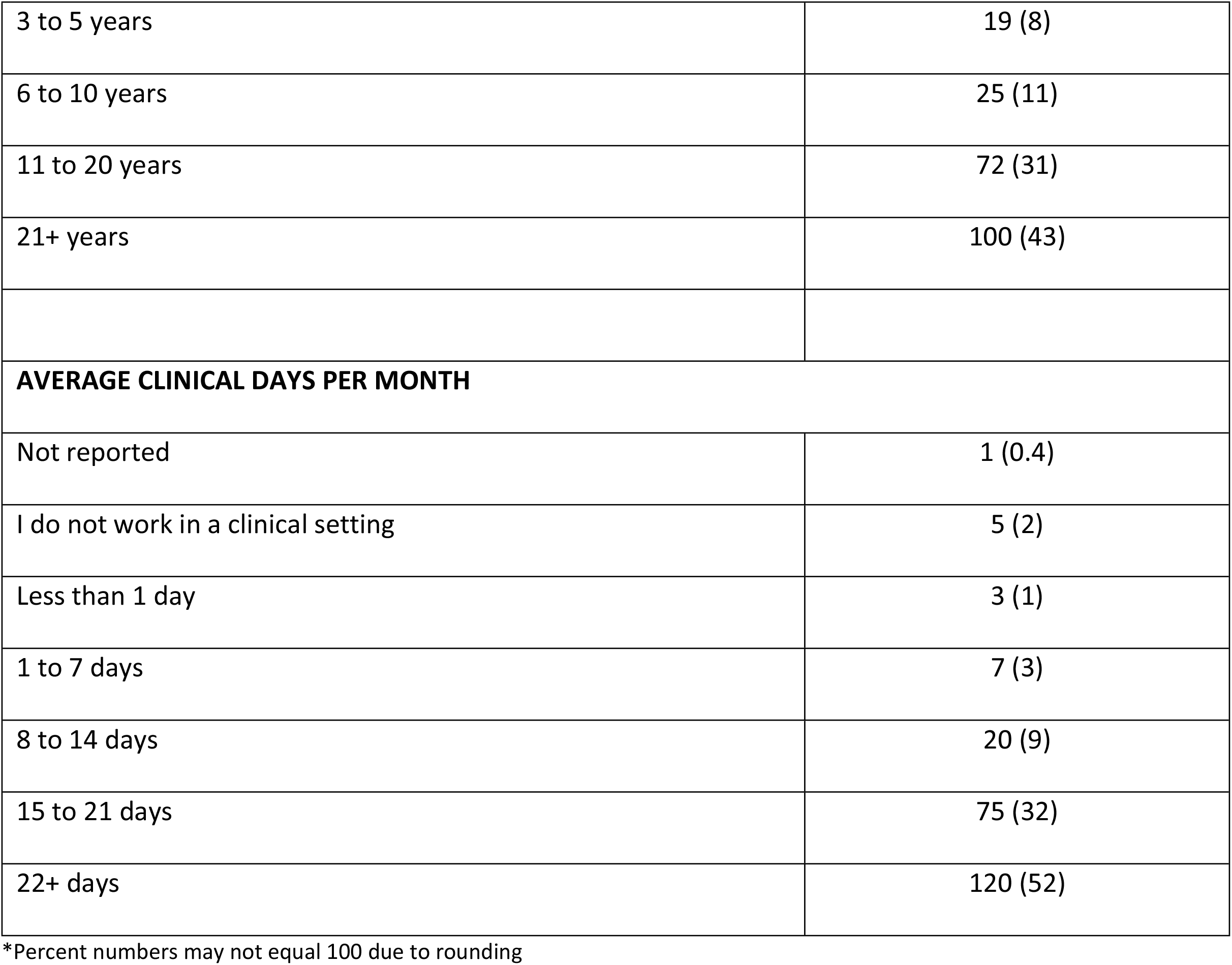
Participant Characteristics

### Physician Quality Improvement Training

#### Physicians With Training

Thirty-one percent of physicians (72 out of 231) received formal training in QI. Within this group of 72 participants, just over half (44 out of 72) identified themselves as at an introductory or intermediate level training (Table 2). Of the 72 individuals trained in QI, 61% (44 out of 72) of respondents ‘agree’ or ‘strongly agree’ that the training prepared them to participate effectively in QI projects. A 5-point Likert scale was used that ranged from ‘strongly agree’ to ‘strongly disagree’ (*M* = 2.68, *SD* = 1). Frequencies for each level are shown in Figure 1.

**Table 2.** Quality Improvement Training

| **QUALITY IMPROVEMENT TRAINING** | **Participants, No. (%\*)** |
| --- | --- |
|  | **(n = 231)** |
| Yes | 72 (31) |
| No | 159 (69) |
| **LEVEL OF TRAINING** | **Participants, No. (%\*)** |
|  | **(n = 72)** |
| Fundamental awareness / Introductory<br>(e.g., understand basic concepts and tools) | 16 (22) |
| Novice (e.g., apply basic tools in small projects) | 28 (39) |
| Intermediate (e.g., certificate program) | 21 (29) |
| Advanced (e.g., formal graduate level training) | 7 (10) |

\*Percent numbers may not equal 100 due to rounding

**Fig 1.**
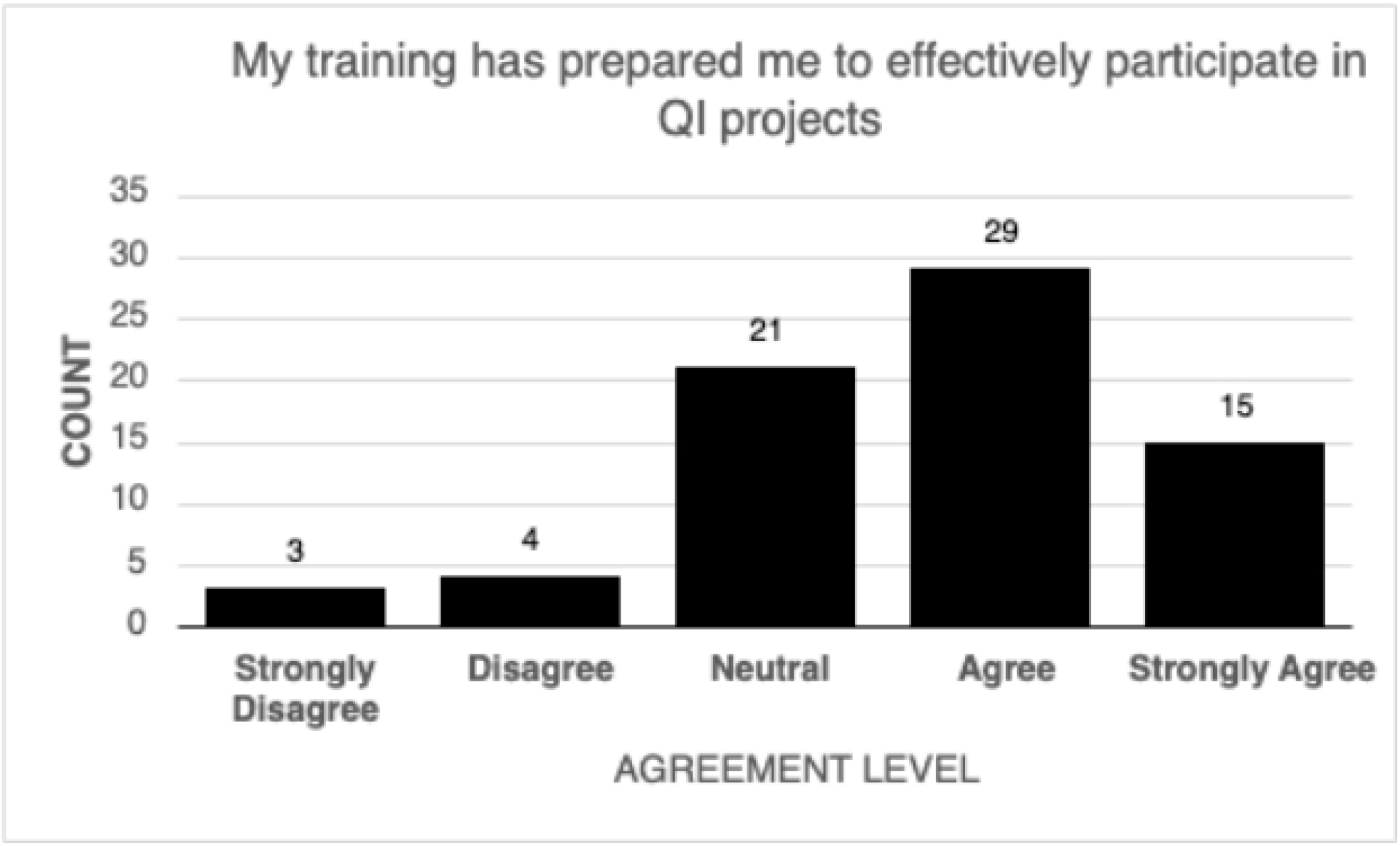
Physician Self-Rating: My training has prepared me to effectively participate in QI projects

The relationship between QI training and years of practice was examined however, the sample sizes within each category were too small to determine a significant effect. Similar findings for the relationship between QI training, interest in receiving QI training, and sex. The differences by sex are negligible and not statistically significant.

#### Physicians Without Training

Of the 159 respondents that did not receive QI training, 60% (96 out of 159) were interested in receiving formal QI training. Respondents were then asked to identify why they were interested in receiving QI training and were able to select multiple responses (Table 3). The top responses were patient-focused, with 88% (84 out of 96) interested in QI training to improve patient care and 86% (83 out of 96) interested in improving patient outcomes. The most frequent responses focused on system and process improvement, with 65% (62 out of 96) interested in improving the efficiency of managerial and clinical processes and 63% (60 out of 96) interested in improving the health system. More than half of respondents who identified an interest in QI training, 53% (51 out of 96), are interested in advancing their clinical skills, and 49% (47 out of 96) would use their training to avoid costs associated with process failures/errors/and poor outcomes.

**Table 3.** Physician Interest in QI Training

| **QI Training** |  |
| --- | --- |
| **REASONS FOR INTEREST IN QI TRAINING †** | **Participants, No. (%\*)**<br>**(n = 96)** |
| Improve care for my patients | 84 (88) |
| Improve patient outcomes | 83 (86) |
| Improve efficiency of managerial and clinical processes | 62 (65) |
| Improve health system | 60 (63) |
| Advance my clinical skills | 51 (53) |
| Avoid costs associated with process failures/errors/and poor outcomes | 47 (49) |
| Advance my career | 33 (34) |
| Meet Ministry of Health accountabilities | 19 (20) |
| Advance my personal knowledge of the topic | 1 (1) |
| Other | 1 (1) |
| **REASONS FOR NO INTEREST IN QI TRAINING †** | **Participants, No. (%\*)** |
|  | **(n = 63)** |
| Not enough time | 31 (49) |
| Too many initiatives going on | 16 (25) |
| Other | 14 (22) |
| Near Retirement | 13 (20) |
| Quality is not effectively measured | 12 (19) |
| I am not interested | 11 (17) |
| Too much bureaucracy | 11 (17) |
| QI not a priority/no need for it | 10 (16) |
| Too expensive/No financial support | 10 (16) |
| I personally get nothing out of it | 9 (14) |
| Lack communication/teamwork | 7 (11) |
| Projects poorly designed/complicated | 7 (11) |
| Leadership not committed | 7 (11) |
| Others (I.e., colleagues) don't participate | 6 (10) |
| Fear negative consequences from leadership | 6 (10) |
| No support (changing practice) | 6 (10) |
| No patient benefit | 6 (10) |
| Clinical setting too small | 6 (10) |
| Resistant to change | 5 (8) |
| Interpretation is not clear | 5 (8) |
| No access | 5 (8) |
| Not invited to participate in design | 5 (8) |
| Not evidence-based | 5 (8) |
| I was never asked to participate | 5 (8) |

† Multiple responses could be chosen by participants

Physicians who had no QI training and indicated they were not interested in training were asked the reasons why they had no interest. More than one response could be selected. Not having the time to participate in training was the top response selected at 49% (31 out of 63). This was cited almost two times as frequently as the second reason, too many initiatives underway at the same time at 25% (16 out of 63).

### Quality Improvement Project Participation

Of the total 231 respondents, 96 had participated in QI projects in the year leading up to the survey. Within this group of 96 participants, 84% (81 out of 96) of the QI projects were at the patient or individual (micro) level, 67% (64 out of 96), were at the organization (meso) level, and 23% (22 out of 96) were at the system (macro) level. Participants were able to select multiple responses. All information related to participation in quality improvement projects is presented in Table 4.

**Table 4.**
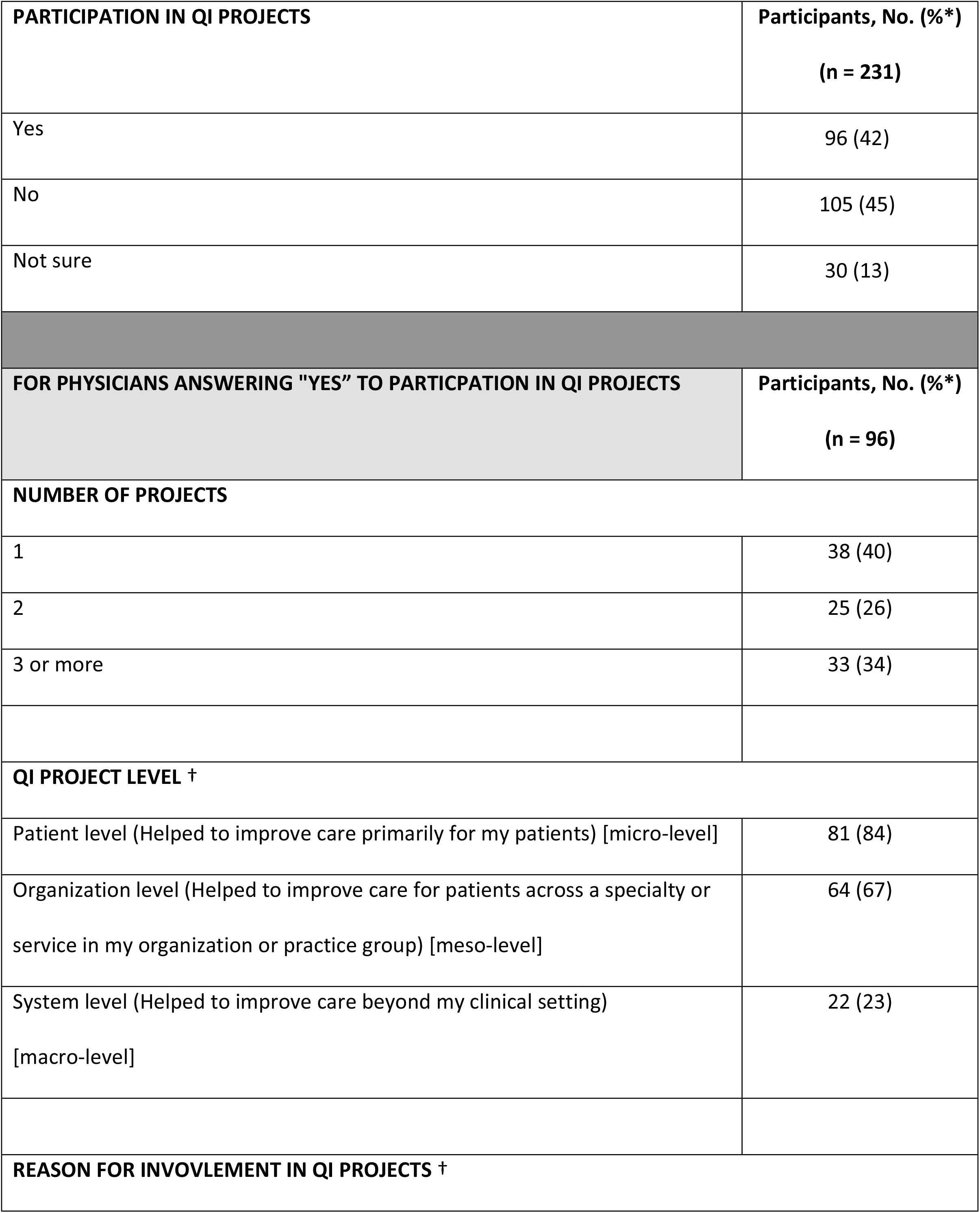
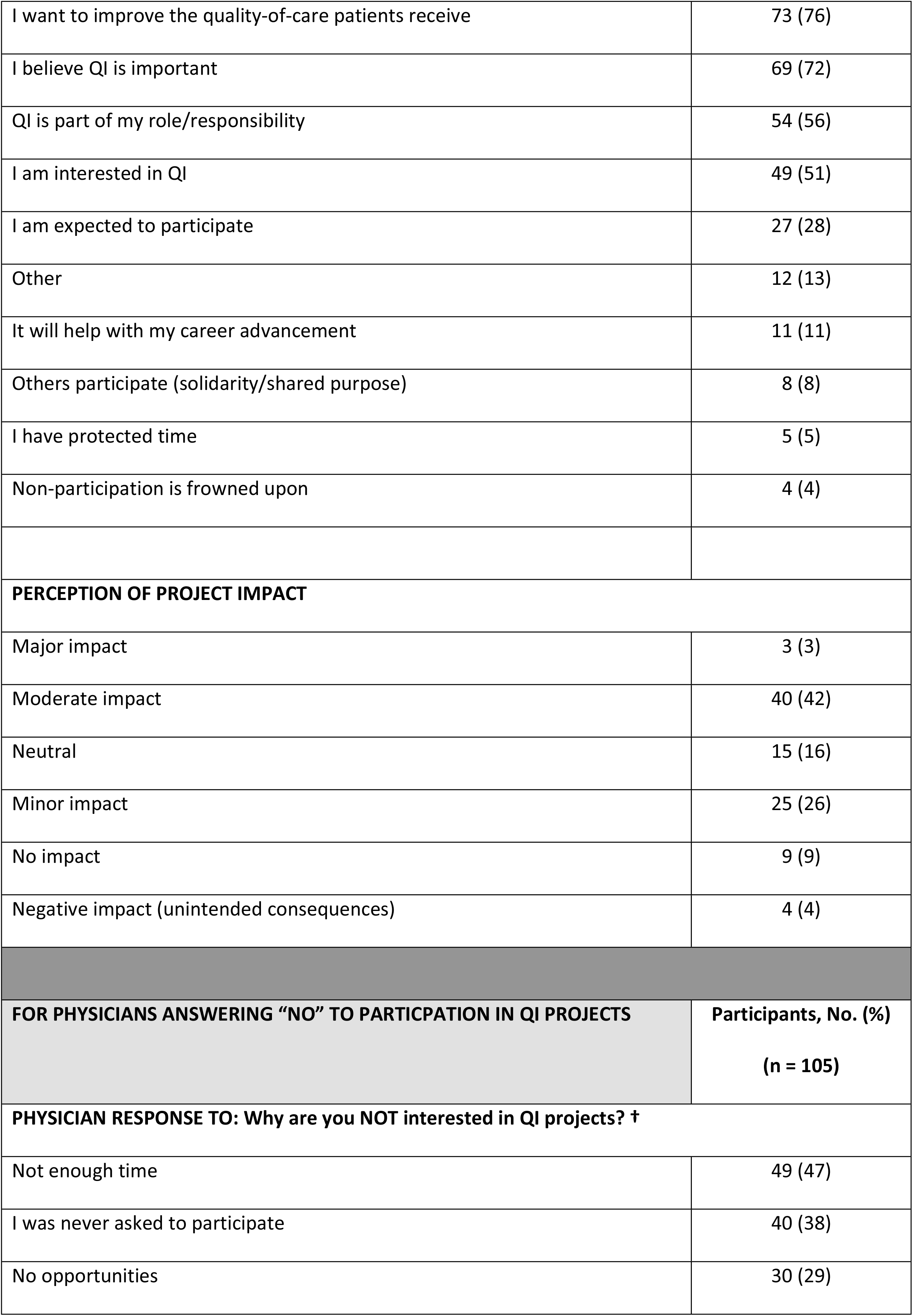
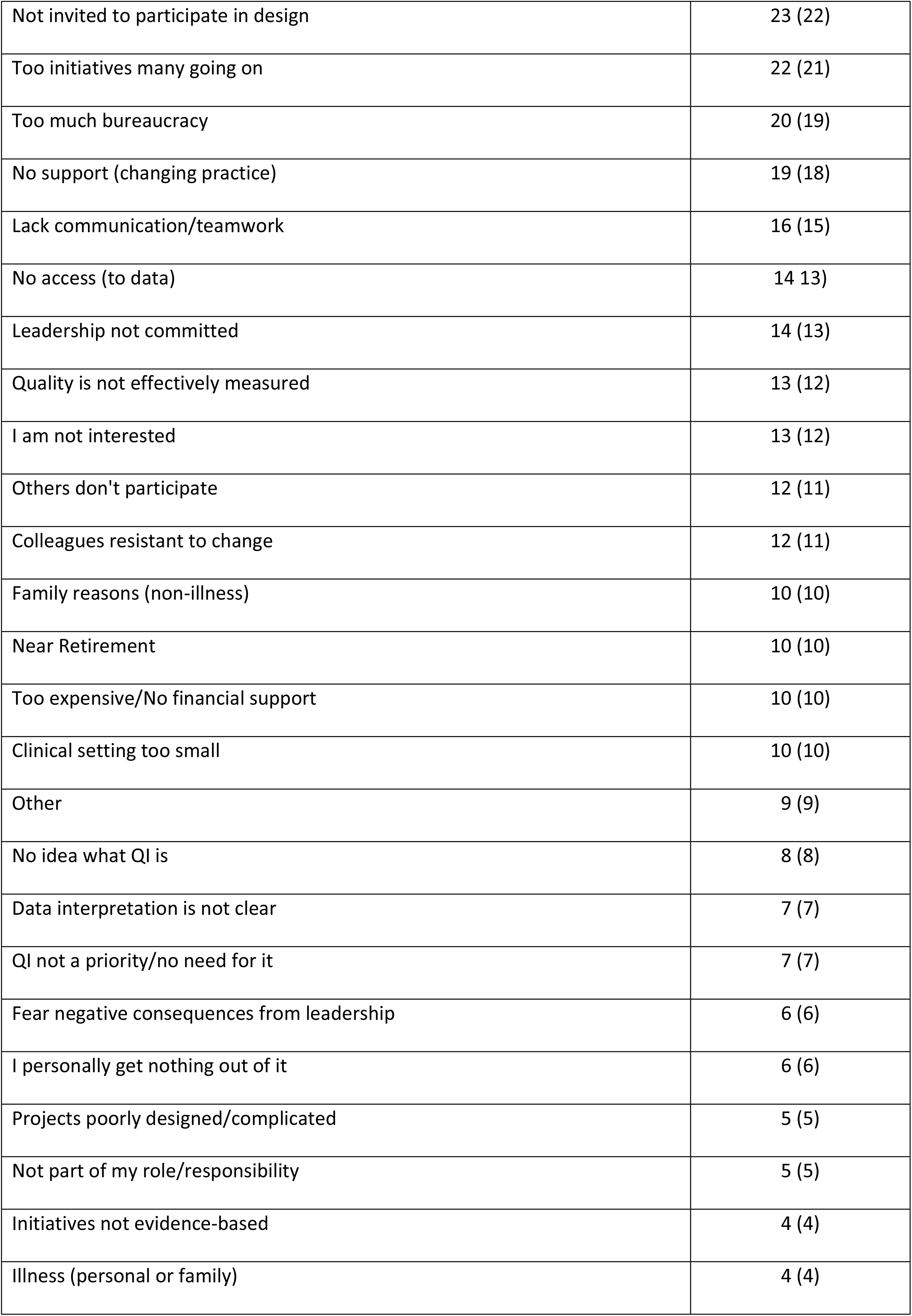
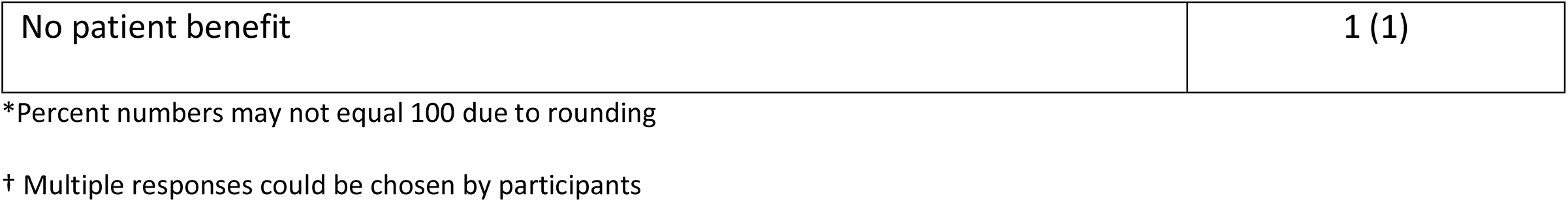
Participation in QI Projects In Past Year

| **PARTICIPATION IN QI PROJECTS** | **Participants, No. (%\*)**<br>**(n = 231)** |
| --- | --- |
| Yes | 96 (42) |
| No | 105 (45) |
| Not sure | 30 (13) |
| **FOR PHYSICIANS ANSWERING "YES" TO PARTICPATION IN QI PROJECTS** | **Participants, No. (%\*)**<br>**(n = 96)** |
| **NUMBER OF PROJECTS** |  |
| 1 | 38 (40) |
| 2 | 25 (26) |
| 3 or more | 33 (34) |
| **QI PROJECT LEVEL †** |  |
| Patient level (Helped to improve care primarily for my patients) [micro-level] | 81 (84) |
| Organization level (Helped to improve care for patients across a specialty or service in my organization or practice group) [meso-level] | 64 (67) |
| System level (Helped to improve care beyond my clinical setting) [macro-level] | 22 (23) |
| **REASON FOR INVOLVEMENT IN QI PROJECTS †** |  |
| I want to improve the quality-of-care patients receive | 73 (76) |
| I believe QI is important | 69 (72) |
| QI is part of my role/responsibility | 54 (56) |
| I am interested in QI | 49 (51) |
| I am expected to participate | 27 (28) |
| Other | 12 (13) |
| It will help with my career advancement | 11 (11) |
| Others participate (solidarity/shared purpose) | 8 (8) |
| I have protected time | 5 (5) |
| Non-participation is frowned upon | 4 (4) |
| **PERCEPTION OF PROJECT IMPACT** |  |
| Major impact | 3 (3) |
| Moderate impact | 40 (42) |
| Neutral | 15 (16) |
| Minor impact | 25 (26) |
| No impact | 9 (9) |
| Negative impact (unintended consequences) | 4 (4) |
| **FOR PHYSICIANS ANSWERING “NO” TO PARTICIPATION IN QI PROJECTS** |  |
|  | **Participants, No. (%)** |
|  | **(n = 105)** |
| **PHYSICIAN RESPONSE TO: Why are you NOT interested in QI projects? †** |  |
| Not enough time | 49 (47) |
| I was never asked to participate | 40 (38) |
| No opportunities | 30 (29) |
| Not invited to participate in design | 23 (22) |
| Too initiatives many going on | 22 (21) |
| Too much bureaucracy | 20 (19) |
| No support (changing practice) | 19 (18) |
| Lack communication/teamwork | 16 (15) |
| No access (to data) | 14 (13) |
| Leadership not committed | 14 (13) |
| Quality is not effectively measured | 13 (12) |
| I am not interested | 13 (12) |
| Others don't participate | 12 (11) |
| Colleagues resistant to change | 12 (11) |
| Family reasons (non-illness) | 10 (10) |
| Near Retirement | 10 (10) |
| Too expensive/No financial support | 10 (10) |
| Clinical setting too small | 10 (10) |
| Other | 9 (9) |
| No idea what QI is | 8 (8) |
| Data interpretation is not clear | 7 (7) |
| QI not a priority/no need for it | 7 (7) |
| Fear negative consequences from leadership | 6 (6) |
| I personally get nothing out of it | 6 (6) |
| Projects poorly designed/complicated | 5 (5) |
| Not part of my role/responsibility | 5 (5) |
| Initiatives not evidence-based | 4 (4) |
| Illness (personal or family) | 4 (4) |
| No patient benefit | 1 (1) |

\*Percent numbers may not equal 100 due to rounding
† Multiple responses could be chosen by participants

Participants were able to choose more than one response when giving the reasons for why they took part in QI projects. The top reasons selected were to improve the quality of care patients received (76%), the belief that QI was important (71%), part of their role/responsibility (56%), and they had personal interest in QI (51%).

Physicians were asked about their perception of the QI projects’ impact on practice in their organizations. Forty-two percent (40 out of 96) thought they had a moderate impact on practice across the organization, however only three percent (3 out of 96) felt they had major impact. Four percent (4 out of 96) thought they had a negative impact with unintended consequences, 9% (9 out of 96) felt they had no impact, 26% (25 out of 96) selected minor impact, and 16% (15 out of 96) indicated a neutral response.

### Organizational Support for Quality Improvement Projects

When asked what the focus was of QI initiatives in their own organizations, half of the physicians (50%) indicated they were unsure. For those that were able to identify the focus of QI projects, safety ranked first (39%), followed by patient-centred (33%), effectiveness (31%), efficiency (28%), timeliness (19%), and equity (12%) (Table 5). Respondents were able to select multiple responses.

**Table 5.** Physicians’ QI Organization Characteristics

| Focus of QI Projects | Participants, No. (%*)<br>(n = 231) |
| --- | --- |
| **PHYSICIAN RESPONSE TO: In my workplace, QI projects focus on making services:†** |  |
| Not sure | 116 (50) |
| Safe | 91 (39) |
| Patient-centered | 76 (33) |
| Effective | 71 (31) |
| Efficient | 65 (28) |
| Timely | 45 (19) |
| Equitable | 27 (12) |

| RESOURCES DEDICATED TO QI (at physician's organization)† | Participants, No. (%*)<br>(n = 53) |
| --- | --- |
| Personnel | 45 (84) |
| Funding | 14 (26) |
| Materials / Equipment | 14 (26) |
| Facilities / Space | 9 (17) |
| Protected Time | 8 (15) |
| Other | 7 (13) |

\*Percent numbers may not equal 100 due to rounding
† Multiple responses could be chosen by participants

A small portion of respondents (53 out of 231) indicated they had dedicated support for QI projects at their workplace. This included personnel (84%), funding (26%), materials/equipment (26%), facilities/space (26%), protected time (15%), and other (13). Respondents were able to select multiple responses.

## DISCUSSION

Physicians outside of hospital settings made up more than half of our participants who completed the study (121 out of 231) and indicated the survey is functional with this group. Overall, participation in our study was low given our recruitment was 231 respondents, and there are 31,500 practicing physicians in Ontario [12]. Despite this, we fulfilled our study objectives conducting a pilot study in preparation for a future large-scale survey. Low response rates are characteristic of studies recruiting physicians for participation [13]. The most important reasons for non-response are lack of time, perceived salience of the study, and concerns about confidentiality [13]. In addition to including an explanation of the value of the research and a clear description of how much time is needed to complete the survey, additional methods such as offering incentives and sending more than one repeated reminder should also be considered. Employing strategies outside of professional association newsletters must be considered for future studies beyond this pilot.

### QI Training and Practice

Our preliminary data provides insight into the QI work, QI training, and perceived barriers for Ontario physicians. We identified a need for robust and applied QI training at the point of care among physicians in this sample. Although two-thirds of our sample reported some QI training, most characterized it as at the introductory or novice level, with less than half describing their training as adequate. Over half of those with no training indicated an interest in receiving training. The first steps to addressing system and organizational demands for efficiency and enhanced effectiveness could be met with targeted in-house QI training.

Physicians not interested in QI training noted they did not have time to participate. Prioritizing and aligning initiatives with organizational strategy could encourage increased QI participation. The positive impact on organizational and system outcomes supported by accurate and effective improvement, has potential to contribute to a high-performing health system [14]

### Barriers to Participating in QI Work

Physicians were asked to complete the phrase, “In my workplace, QI projects focus on making services [*fill in the blank*]” by selecting one of the National Academy of Medicine’s (formerly the Institute of Medicine) six domains of quality [15] which were listed as: safe, patient-centred, effective, efficient, timely, and equitable. Half of the respondents answered they were ‘not sure’, indicating the need for unambiguous objectives, active engagement of relevant stakeholders, and clear and regular communication about QI initiatives. ‘Equitable’ was chosen least often by our respondents and suggests that organizations may want to devote time and resources to raising awareness of the relevance and importance of this quality domain in delivering high quality care.

A small group of physicians indicated they had some support for QI work, with protected time reported least often. Lack of protected time impacts QI work in organizations. A recent study by Deilkås and colleagues [6] reported physicians wanted to participate in QI work, but few had designated time for this activity. As well, physicians with designated time participated significantly more [6].

### Limitations

The low response rate plus the sample size meant it is likely not all groups are represented with regard to demographics. The newsletters of the Ontario Medical Association (OMA) and the Ontario Hospital Association (OHA) were used to contact physicians for recruitment and between the two, they have the potential to provide a connection to all practicing physicians in Ontario. However, the current policies of both organizations prevent research surveys from being sent directly to physicians from a research team. It was necessary to test this recruitment strategy, but other approaches will have to be considered in the future. Also, the study was conducted during the COVID pandemic, and there is a high potential that the overall physician burden may have impacted response rates.

### Future Research

Our preliminary results suggest training initiatives may need attention including looking at standards and solutions for training for those that are interested in participating. Our pilot study provides the data needed to move forward in conducting a full-scale survey that would provide more robust results.

## CONCLUSION

Physician involvement in QI contributes to the success and sustainability of these QI initiatives. The first step is to accurately measure physicians’ skill set using the survey we developed. Before launching a full-scale survey, it is necessary to assess the feasibility of processes, time, and resources to identify challenges. We identified the need for an extensive recruitment strategy and attention to the timing of conducting the survey.

## Data Availability

Data are available from the OHA for researchers who meet the criteria for access to confidential data.

## Acknowledgements

We are grateful to all physicians for completing and submitting survey responses. We thank the Ontario Medical Association and Ontario Hospital Association for assistance with recruitment. We thank Ontario Health for there assistance developing the survey.

## Data Availability

The data that support the findings of this study are available from the corresponding author upon reasonable request.

## SUPPORTING INFORMATION

S1 Appendix. Physician speciality. (DOC)

